# A Pilot Validation Study Comparing FIBI, a Slide-Free Imaging Method, with Standard FFPE H&E Tissue Section Histology for Primary Surgical Pathology Diagnosis

**DOI:** 10.1101/2022.03.10.22272226

**Authors:** Alexander D. Borowsky, Richard M. Levenson, Allen M. Gown, Taryn Morningstar, Thomas A. Fleury, Gregory Henderson, Kurt Schaberg, Amelia B. Sybenga, Eric. F. Glassy, Sandra L. Taylor, Farzad Fereidouni

## Abstract

**Introduction:** Digital pathology whole slide images (WSI) have been recently approved by the FDA for primary diagnosis in clinical surgical pathology practices. These WSI are generated by digitally scanning standard formalin-fixed and paraffin-embedded (FFPE) H&E-stained tissue sections mounted on glass microscope slides. Novel imaging methods are being developed that can capture the surface of tissue without requiring prior fixation, paraffin embedding, or tissue sectioning. One of these methods, FIBI (Fluorescence Imitating Brightfield Imaging), an optically simple and low-cost technique, was developed by our team and used in this study.

**Methods:** 100 de-identified surgical pathology samples were obtained from the UC Davis Health Pathology Laboratory. Samples were first digitally imaged by FIBI, and then embedded in paraffin, sectioned at 4 µm, mounted on glass slides, H&E stained, and scanned using the Aperio/Leica AT2 scanner. The resulting digital images from both FIBI and H&E scan sets were uploaded to PathPresenter and viewed in random order and modality (FIBI or H&E) by each of 4 reading pathologists. After a 30-day washout, the same 100 cases, in random order, were presented in the alternate modality to what was first shown, to the same 4 reading pathologists. The data set consisted, therefore, of 100 reference diagnoses and 800 study pathologist reads (400 FIBI and 400 H&E). Each study read was compared to the reference diagnosis for that case, and also compared to that reader’s diagnosis across both modalities for each case. Categories of concordance, minor and major discordance were adjudicated by the study team based on established criteria.

**Results:** The combined category, concordance or minor discordance, was scored as “no major discordance.” The overall agreement rate (compared to the reference diagnosis), across 800 reads, was 97.9%. This consisted of 400 FIBI reads at 97.0% vs. reference and 400 H&E reads vs. reference at 98.8%. Minor discordances (defined as alternative diagnoses without clinical treatment or outcome implications) were 6.1% overall, 7.2% for FIBI and 5.0% for HE.

**Conclusions:** Pathologists without specific experience or training in FIBI imaging interpretation can provide accurate diagnosis from FIBI slide-free images. Concordance/discordance rates are similar to published rates for comparisons of WSI to standard light microscopy of glass slides for primary diagnosis that led to FDA approval. The present study was more limited in scope but suggests that a follow-on formal clinical trial is feasible. It may be possible, therefore, to develop a slide-free, non-destructive approach for primary pathology diagnosis. Such a method promises improved speed, reduced cost, and better conservation of tissue for advanced ancillary studies.

## Introduction

While in-vivo methods for disease detection and classification, including radiology and liquid biopsy, continue to improve with respect to spatial resolution (radiology) and molecular information, they have not, to date, obviated the need for tissue biopsy and/or excision, methods that remain the current standard for all cancer and many non-cancer diagnoses in clinical medicine [1, 2]. However, tissue processing of these biopsies incurs a significant delay, typically overnight, and not infrequently some tissue is lost or discarded in this process. The resulting standard tissue sections mounted on glass slides can then be scanned using automated whole-slide microscopy systems, and then reviewed on computer displays; this approach has recently been approved by the FDA to support primary surgical pathology diagnosis in clinical medicine [3-5]. However, while such digital imaging comes with real advantages, including being a necessary pre-requisite for computational pathology approaches, they do not solve some major logistical challenges posed by conventional pathology, and in fact contribute to them. This is because the time-and effort-consuming workflow involved in standard slide production is still required, and in fact, the scanning step, which occurs after the slides are produced, represents another costly and time-consuming step.

It would be desirable if histology information could be acquired even before slides are generated, cutting hours and expense from the diagnostic process. The current approach for rapid histology involves intraoperative assessment using frozen sections; the process can be performed quickly, but requires skilled input, consumes potentially valuable tissue, and often generates substandard histology results due to artifacts associated with tissue freezing and other steps in the cryotomy process.

In response to a perceived need for rapid, slide-free histology a number of tissue imaging technologies are under development, but adoption to date has been primarily in research-only applications. These approaches include microscopy with UV excitation (MUSE) [6], light-sheet microscopy [7, 8], structured illumination microscopy [9], optical coherence microscopy [10], stimulated Raman histology [11], and most recently, photoacoustic-based systems [12], all useful and promising approaches, with current implementations presenting some challenges with respect to cost, resolution and/or throughput.

Here, we describe a new method for rapid slide-free histology, termed FIBI (Fluorescence Imitating Brightfield Imaging). FIBI is based on the use of absorbing (not necessarily fluorescing) stains applied briefly to the specimen, coupled with 405 nm illumination. The stains penetrate the surface of the tissue to approximately 10 μm (data not shown), limiting in-focus image creation to approximately that depth. The 405 nm excitation light generates broad-spectrum autofluorescence diffusely inside the bulk of the thick (unsectioned) sample. This generated autofluorescence then back-illuminates the stained layer to generate an image of the surface layer of the tissue. Conveniently, stains familiar to histopathologists, namely, hematoxylin and eosin (H&E), perform well in this application. Hematoxylin is a strongly absorbing, non-fluorescing dye that binds predominantly to nuclei. Eosin staining provides a red-pink tint to protein and other non-nucleic acid elements, just as it does in conventional slide-based histology, while contributing additional contrast due to its intrinsic fluorescence [13].An example of a FIBI image of benign breast is shown in Figure 1.

**Fig. 1.**
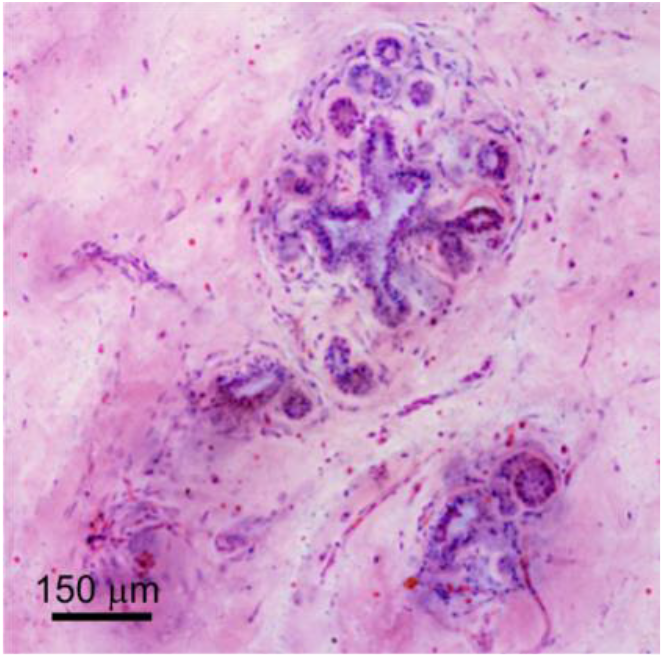
FIBI image of benign breast lobules, ducts, stroma and microvasculature.

**Fig. 2.**
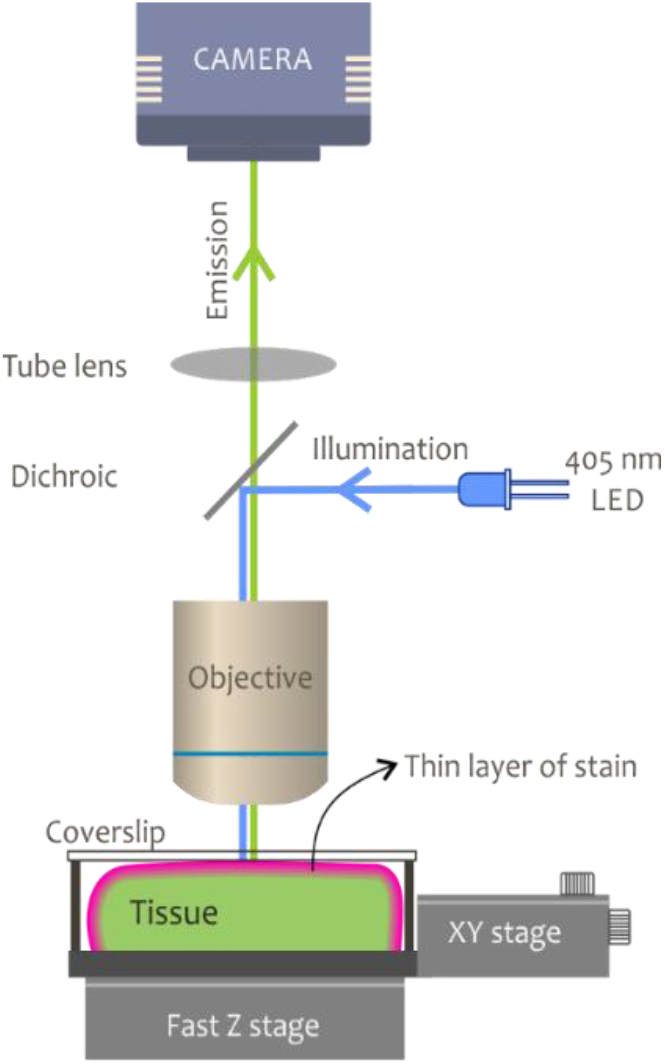
Schematic design of a FIBI imaging microscope.

This report presents a comparison study of pathologist diagnoses using FIBI technology over a variety of surgical pathology specimens (See Table 1) compared to standard whole-slide images (WSI) of FFPE H&E 4-µm tissue sections.

**Table 1:** Case distribution by primary tissue type

| Organ | Number of Specimens |
| --- | --- |
| Adrenal Gland | 3 |
| Appendix | 4 |
| Bladder | 3 |
| Breast | 5 |
| Colon | 8 |
| Kidney | 7 |
| Larynx | 1 |
| Liver | 2 |
| Lung | 5 |
| Ovary | 5 |
| Pancreas | 7 |
| Parotid Gland | 3 |
| Prostate | 10 |
| Salivary Gland | 1 |
| Seminal Vesicle | 1 |
| Skin | 5 |
| Small Bowel | 2 |
| Soft Tissue | 1 |
| Stomach | 3 |
| Testis | 1 |
| Thyroid | 6 |
| Tongue | 5 |
| Tonsil | 3 |
| Uterus | 9 |
| **Total** | **100** |

**Table 2.** Results summary.

|  | CONCORDANT | MINOR DISC. | MAJOR DISC. |
| --- | --- | --- | --- |
| H&E WSI | 375 (93.8%) | 20 (5%) | 5 (1.2%) |
| **FIBI** | **359 (89.8%)** | **29 (7.2%)** | **12 (3.0%)** |
| Total | 734 (91.8%) | 49 (6.1%) | 17 (2.1%) |

## Methods

### FIBI imaging instrumentation and procedure

Figure 1 presents a schematic drawing of the instrumentation. The tissue sample are flattened against a glass coverslip inside a custom-built specimen holder with an imaging window of 25 × 35 mm^2^. Illumination is performed using a 405 nm LED (LZ1-00UB00, LED Engin) directed to the sample via a broadband dichroic (Di03-R405-t1-25x36, Semrock). The scanning is performed with an average speed of 1 cm^2^/min with XY travel range of up to 25 × 50 mm^2^, along with a Z-positioner (Zaber Technologies, Vancouver, Canada) to provide focus stacking. A 10X, NA=0.3 Plan Fluor objective (Nikon, Japan) was employed for this study, although both 10X NA=0.45 and 20X objectives can also be used, and images were collected with a 9-megapixel scientific-grade CCD color camera (Ximea, MD091CU-SY) connected via a 160-mm tube lens (Thorlabs TTL 165-A).

Images were acquired using a focus stacking technique to create an extended depth of field (EDOF) images required to accommodate slight tissue surface unevenness [14]. The captured images were color-corrected using a custom histogram-matching tool [15] and processed to improve contrast and sharpness using the AForge.NET open-source library. Finally, the images were stitched and pyramidal svs files created using a stitching library from Microvisioneer (Esslingen am Neckar, Germany). Image acquisition, alternation between light sources, stage movement and focusing was performed using control software developed in the .NET environment.

In order to determine if the image quality obtained using FIBI was non-inferior to conventional FFPE H&E WSI we designed a study comprising 100 surgical pathology cases, and recruited 4 reading pathologists (TF, GH, KS, AS) who had no prior experience viewing FIBI-acquired images. They were asked to provide a diagnosis for each case twice, seeing either the FIBI case first or the H&E case first, and then, after a 30-day washout period, they were presented with the same specimens imaged with the alternative technique. That is, for some cases they saw the FIBI (or H&E) images first, and then, 30-days later, the H&E (or FIBI) images of the same cases. All 100 specimens were read by all study pathologists in both modalities. We compared the concordances of diagnoses for these 800 reads to the reference diagnoses that had been determined by two experienced pathologists (AMG, ADB) who were not part of the reading panel. The results were then interpreted in line with analyses used to evaluate performance in consensus recommendations used for WSI vs. glass slide comparison studies performed as part of the FDA approval process used in the Leica/Aperio scanner submission[5].

### Specimens

100 surgical pathology specimens from the UC Davis Health clinical laboratory in surgical pathology were chosen to span common specimen types from available excess tissues not needed for the initial diagnosis and designated for disposal. In all cases the tissues used were fixed in formalin, but not otherwise processed. Tissues used for the study were all obtained without protected health information (i.e., deidentified) and were obtained under a UC Davis IRB determination of “not-human subjects research.” Selection from this cohort biased the cases toward excisions, as much smaller biopsy specimens are typically processed in their entirety (and therefore would not have been available for this study). In addition, because the most morphologically relevant tissue regions may have been mostly or entirely processed as part of the clinical workup, residual discarded tissue might consist only of adjacent benign material. This yielded an array of tissue types and diagnoses including normal/benign, hyperplasias, benign neoplasms, cancers, and various inflammatory lesions. See Table 1 for sites of origin; a complete list of reference diagnoses is provided in Supplementary Table 1.

### FIBI imaging

In preparation for imaging, the tissue available for each case was examined in conjunction with the de-identified pathology report, and a selected area corresponding to the size of a typical medium-to-large specimen (i.e., up to about 2.0 × 2.5 cm^2^) was manually dissected using a razor blade to achieve a tissue surface as flat as possible.

FIBI imaging was performed by first staining the tissue with 0.5 mg/ml Mayer’s hematoxylin (H3136, MilliporeSigma) for 30 seconds. The tissue was then rinsed in deionized (DI) water before staining with 0.25% eosin Y (E4382, MilliporeSigma) in 79% ethanol for 30 seconds. Following staining, the tissue was rinsed twice with DI water for 30 seconds. The tissue was placed in a holder designed to gently compress the specimen against an imaging coverslip, and the whole tissue specimen was scanned with the FIBI instrument described above, with EDOF enabled.

### H&E imaging and case curation

After FIBI imaging, the tissue sample was then submitted for standard histology involving paraffin embedding and microtome sectioning at 4 µm, mounting on glass slides, H&E staining, and coverslipping; the resulting slides were then scanned on an Aperio/Leica AT2 slide scanner at 20X. The resulting FIBI and H&E WSI were uploaded to PathPresenter (www.pathpresenter.net) for viewing. The supervising study pathologist (ADB) ensured that each image captured the study reference diagnosis. In some cases, this reflected the actual clinical diagnosis (obtained from the de-identified pathology report), but if the study pathologist determined that the sections included in the study series did not contain tissue regions that reflected that diagnosis, an alternative reference diagnosis was recorded. For example, the study tissue may have been comprised of benign tissue even if the primary case diagnosis recorded the presence of cancer. Minimal clinical data was provided to the reviewing pathologists, consisting of patient age and sex and tissue sampling origin. If, in the opinion of the lead study pathologist, additional clinical metadata was needed to arrive at the reference diagnosis (as would be available in a true clinical context), this was added to the available metadata.

### Case reading methodology

Four “reader” pathologists were recruited to the study. All are board certified in Anatomic Pathology, with varying experience in general and specific surgical pathology (gastrointestinal: KS; pediatric: ABS), with two in academic and two in private surgical pathology practice.

Case order and modality (FIBI versus H&E) was randomized for each pathologist to view only those images on PathPresenter, a cross-platform digital image viewer. (Note that in the discussion that follows, for convenience and familiarity, we refer to the conventionally sectioned slide-based images as “H&E” even though the thick (FIBI) and thin-cut specimens were both stained with hematoxylin and eosin.) The study pathologists were asked to provide the best/most likely diagnosis they could arrive at without additional information or ancillary studies such as IHC and were also asked to comment about adequacy. They were advised that any resource, textbook, medical literature, online database that they would use clinically to help arrive at the best diagnosis was permitted. They were not permitted, however, to consult with a colleague even though this is common in clinical practice. If they would obtain IHC in clinical practice, or if they would send a case for expert consultation, they could add a note or comment, but these steps were not permitted in the study.

After a 30-day washout period, the same 100 cases were again provided—but with the modality switched for each case. If the observer saw the H&E for a case in the first set, in the second round they had access only to the FIBI image, and vice versa. They were not able to view their prior diagnoses or the previous set of slides.

### Data collection and adjudication

The diagnoses were compared to the reference and scored as concordant, minorly discordant or majorly discordant. “Minor discordance” was defined as a difference in the diagnosis that would not lead to a different clinical treatment or outcome, whereas “major discordance” implies that the difference in the diagnosis would lead to a different treatment recommendation or outcome [5]. All diagnoses and concordance scores were reviewed independently by two pathologists (ADB and AMG), and any differences in concordance assignment were discussed to reach a consensus.

For the purposes of the primary endpoint, major discordance rates were compared to the combined rates of concordance and minor discordance. In addition, intraobserver variation was determined by comparing a reading pathologist’s diagnosis from one modality to the other. In this way, although a reader may have been in disagreement with the reference diagnosis, discrepancies due solely to the impact of the modality used could be assessed.

To determine if the FIBI method was non-inferior to H&E, a binomial regression with an identity link was fit to model the percentage of major discordance versus method. Because observations by the same pathologist and on the same specimen were correlated, we estimated model parameters with generalized estimating equations and used a robust covariance matrix to estimate the variance. The FIBI method was deemed to be non-inferior to H&E if the upper bound of a one-sided 95% confidence limit for the difference between the FIBI and H&E method was less than the prespecified non-inferiority margin of 4.0%. PROC GENMOD in SAS Version 9.4 was used to fit this model.

## Results

Images were acquired directly from the surface of intact fixed specimens, which were then submitted for standard histology, attention being paid to having the same face of the tissue available for sectioning and staining that was previously captured during the original FIBI scan. An example of a whole specimen image, similar to what was presented to the reviewers via PathPresenter, is shown in Fig. 3. Reviewers had control over orientation (rotation) and zooming, as well as simple image display parameters (brightness and contrast).

**Fig. 3.**
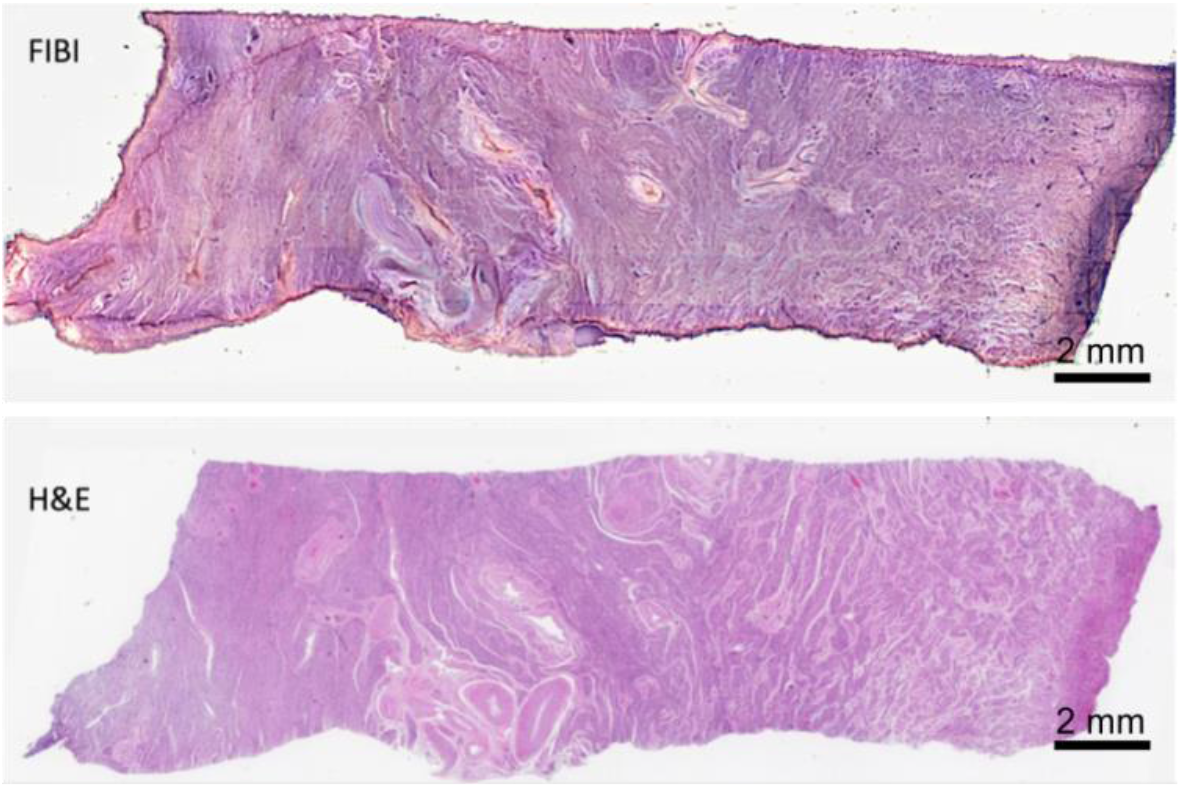
Cervix specimen imaged via FIBI (top) and after H&E slide preparation and WSI scanning, bottom. Screen shots from PathPresenter

The distribution of tissues of origin of the cases in the study set is shown in Table 1 and diagnoses listed in Supplemental Data section Table 1. Overall, 54% were malignant, 26% showed non-malignancy-related pathology (e.g., proliferative, inflammatory), and 20% showed no evidence of disease.

### Concordance

Determination of concordance was performed by comparing reader diagnoses to <u>reference diagnoses</u>, and also by comparing the reader diagnosis from FIBI to the <u>same-reader diagnosis</u> from H&E. Adjudication followed criteria established with FDA guidance and used in similar studies [5]. The rates of concordance, minor discordance and major discordance were tabulated for each reader pathologist, and for each image modality.

Over the 800 reader diagnoses, the overall rates for both modalities combined was 91.8% concordant, 6.1% minor discordance, and 2.1% major discordance. The rates for H&E-only were: 93.8% concordant, 5.0% minor discordance, and 1.2% major discordance. The rates for FIBI-only were: 89.8% concordant, 7.2% minor discordance, and 3.0% major discordance. The difference (delta) in the major discordance rates between FIBI and H&E was 1.8% with a one-sided upper 95% confidence interval of 3.1%, meeting the predetermined criterion for acceptance of less than 4.0%.

Intraobserver discrepancies were assessed by first comparing each reader’s H&E-to-FIBI diagnosis. Considering the relative contribution of discordances in each modality, versus identical discordance in both modalities for a given case from a given observer, we audited all major discordances with respect to the reference diagnosis to determine if the discordances were modality-specific. For all readers combined, the H&E-to-FIBI rate of intraobserver no-major-discordance was 98.2% (1.8% or 14 of 17 major discordances) with 4.0% minor differences. Of the H&E majorly discordant diagnoses (N=5), three out of five had the same discordant diagnosis from that reader using FIBI (60% internal concordance in the H&E-to-FIBI direction). Of the FIBI major-discordant diagnoses (N=12) three of 12 had the same discordant diagnosis from that reader using H&E (25% internal concordance in the FIBI to H&E direction).

### Example image pairs

A pair of FIBI and corresponding H&E images from a benign breast specimen is shown in Figure 4. Even at low digital magnification, the lobules are clearly visible in both presentations. In general, looking at higher power, there is a close resemblance between the modes, but there are discernable differences in image properties. FIBI images can display contrast not in evidence in standard H&Es. For example, lipid is still present (and imageable) in the intact tissues imaged with FIBI, but as lipids are removed during the solvent-based paraffin-embedding process, lipid within adipose regions becomes empty “white” areas on H&E-stained slides, as can be seen in the lower areas of whole specimen images (top row), which also highlights adipose blood capillaries presence not readily visible in the H&E version.

**Fig. 4.**
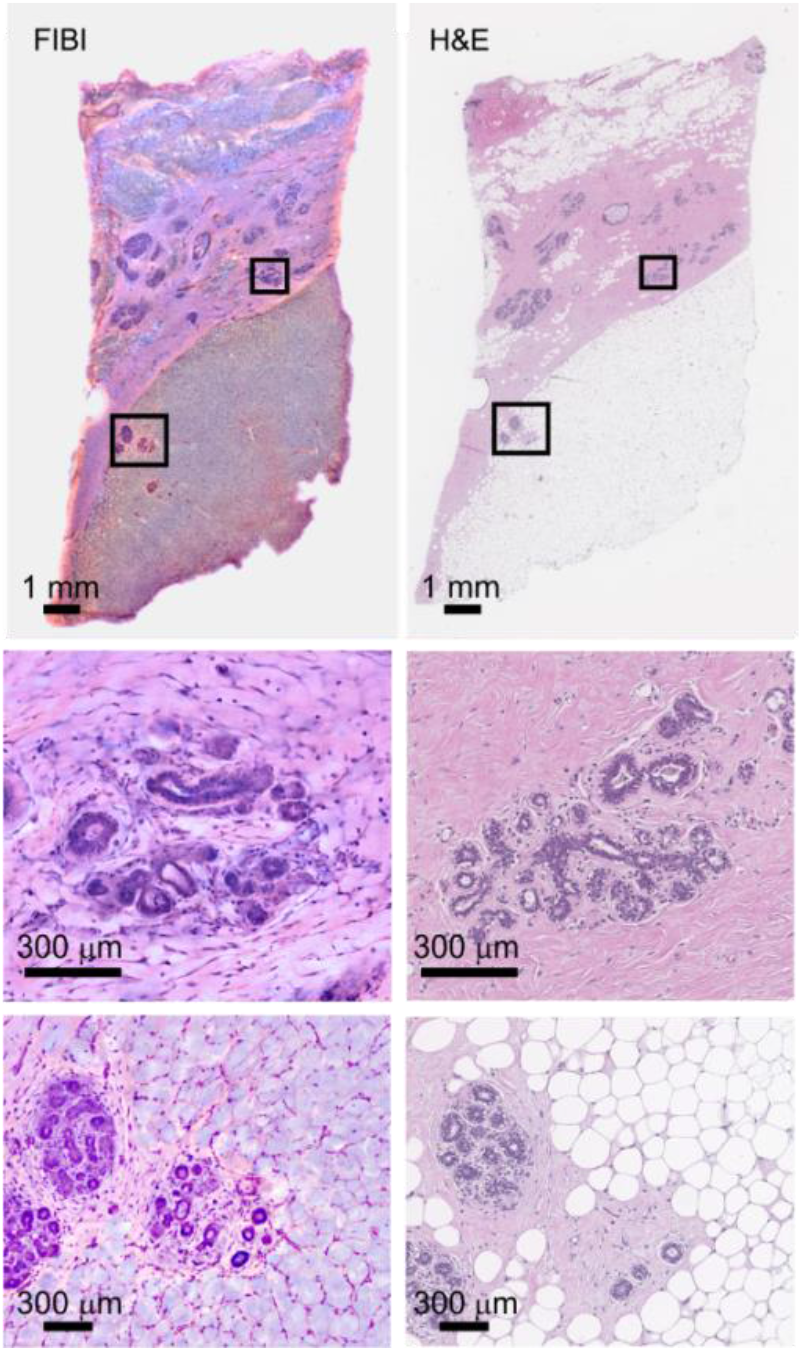
Normal breast. Left, FIBI, Right H&E. Closeups of a typical lobule are shown in the middle row. Bottom row: Lipid and capillaries in adipose tissue are visible in the FIBI specimen.

Figure 5 illustrates a case of invasive lobular carcinoma imaged via FIBI (left) and H&E (right). Zoomed-in regions in each mode show a duct surrounded by invasive lobular carcinoma cells.

**Fig. 5.**
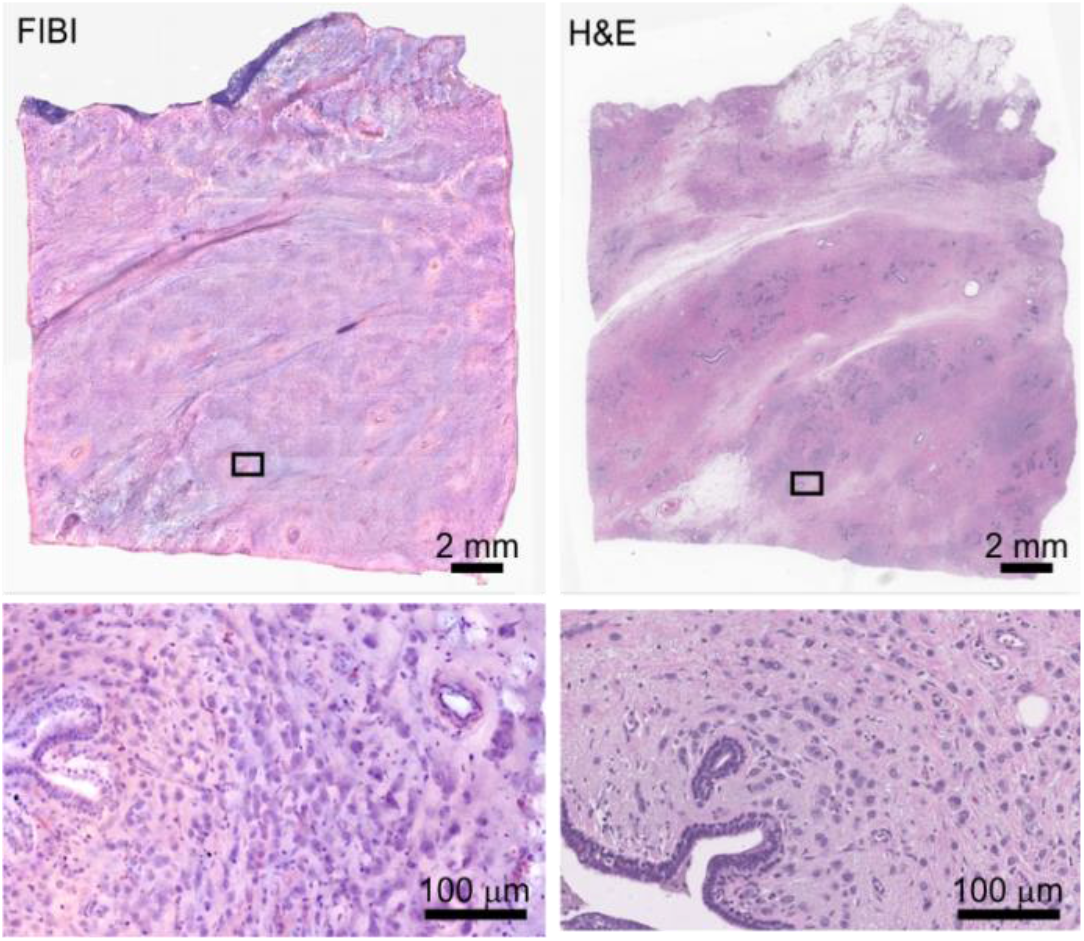
FIBI and corresponding H&E images of breast with invasive lobular carcinoma. Whole specimen (top) and zoomed-in regions (bottom) showing ducts surrounded by lobular carcinoma cells.

An example of a tongue squamous cell carcinoma specimen exhibiting invasive squamous carcinoma along with necrosis is present in Fig. 6. Resolution in the FIBI image is sufficient to demonstrate chromatin appearance and the presence of mitoses, as shown in the lower row in both the FIBI and H&E fields.

**Fig. 6.**
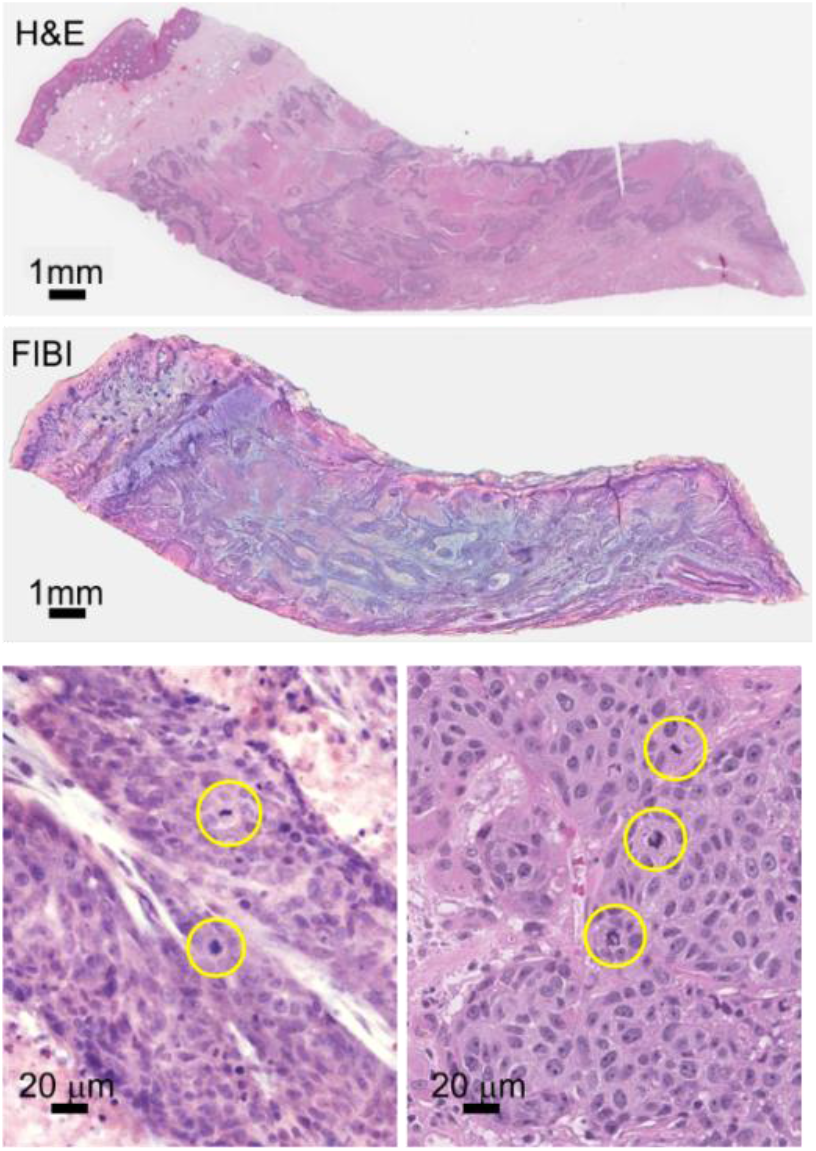
Tongue squamous cell carcinoma with invasive squamous carcinoma and necrosis. Lower row: mitoses are visible in both FIBI and H&E images.

## Discussion

For more than a century diagnostic pathology has relied upon bright-field transillumination microscopy of thinly sliced and stained tissues. The preparation of these specimens for microscopic analysis entails a sequence of steps (fixation, dehydration, infusion with paraffin, oriented block mounting, microtome sectioning at 4 µm, transfer to a glass slide, rehydrating and staining with histology dyes, and coverslip mounting). In aggregate, this process is time- and resource-demanding.

Digital pathology, the analysis and evaluation of tissue sections that have been converted to a digital image using commercial slide scanners and then examined and evaluated on a computer or monitor screen, has several theoretical advantages over standard analog pathology based on direct viewing of glass slides. Some of these advantages include the ability to share images instantaneously with other pathologists in different locations, and, when coupled with image analysis and machine learning, the potential for greater accuracy, reproducibility and standardization of pathology diagnoses and prediction of patient outcome [16].

Two large seminal studies conclusively demonstrated the “non-inferiority” of digital-image-based diagnoses compared with microscopic slide evaluation [5, 17]. In the study by Mukhopadhaya et al. [17], 1992 cases were studied, corresponding to 15,925 reads, the major discordance rate with the reference standard diagnosis was 4.9% for digital images and 4.6% for microscopy. The overall major discrepancy rate with respect to reference standard diagnoses in [5], in which 2045 cases and 5849 slides were examined, was 3.6% for digital images, and 3.2% for manual slide review.

In this report, we have described FIBI, a novel slide-free digital histology system that permits rapid acquisition of digital images, taking minutes instead of hours or days as required by traditional slide-based pathology followed by whole-slide imaging. Furthermore, we have demonstrated the ability of pathologists to render accurate diagnoses base on their evaluation of these FIBI-acquired digital images, with an accuracy within the range of existing standard histology and WSI systems. Results of the present study examining the suitability of FIBI-enabled histology demonstrated an accuracy of 97.0%, well within the ranges reported for conventional WSI approaches.

The development of FIBI comes at a time when advances in optics, light sensors, microcomputer power, and algorithmic methodologies have dramatically increased the available strategies for optical microscopy in recent years. But while technology may permit development of slide-free optical microscopy, any compelling slide-free microscopy system that has potential for introduction into diagnostic surgical pathology laboratories ideally should be based on simple and affordable hardware, produce images within minutes not hours, not be destructive of tissue, generate a wide field of view (FOV), and be able to produce images that closely simulate those of scanned-in images of conventional slide-based hematoxylin and eosin (H&E) stained tissue, permitting accurate diagnostic assessment by the pathologist.

A recent review highlights a number of slide-free histopathology imaging systems [18], some of which can produce images that approximate the appearance of slide-based hematoxylin and eosin (H&E)-stained tissue. These include confocal microscopy [19], nonlinear microscopy [20], structured illumination microscopy phase detection [21], microscopy with UV surface excitation (MUSE) [6, 11], full-field optical coherence tomography [22], light-sheet microscopy [23, 24], and photoacoustic microscopy [12]. Among this group, some approaches have an ability to generate contrast without requiring exogenous stains (for example, multiphoton [25] and photoacoustic techniques [12]) or to create 3-dimensional data that provide information beyond what is visible on 2-dimensional thin sections (for example, light sheet systems [24]). However, these methods can involve relatively complex optics, large data set generation and computationally dependent image display.

Among the slide-free approaches, MUSE and FIBI are differentiated because of their use of simple, affordable hardware that produce images that pathologists can interpret much as they would standard H&E-stained tissue sections. FIBI differs from MUSE mostly in that the contrast used to generate the images is based on staining with the familiar dyes, hematoxylin and eosin; induced broad-band “virtual” backlighting then generates images that directly resemble standard histopathology—no image intensity inversion or major re-coloring steps are required. Moreover, FIBI offers an order of magnitude faster imaging rate, due to achievable high brightness, and because of epi-illumination (through-the-lens excitation), it is straightforward to include an objective turret with a variety of lenses with different magnifications. A current limitation of FIBI is that the acquired images resemble standard microscopy of physical sections. So far, the optics produce images that would be directly comparable to a slightly thicker FFPE section, perhaps 6-8 μm. Comparing to a 4 μm standard section, about 30% more nuclei can be seen in dense cellular areas (data not shown). Optical and computational methods to further “pseudo-thin” the FIBI images are feasible in future implementations.

In this pilot validation study, we have demonstrated the ability of FIBI to produce direct-to-digital images within minutes that permit pathologist assessment with an accuracy well within CAP and FDA guidance for approvals of digital pathology WSI for primary diagnosis [4, 5]. In contrast to traditional WSI digital pathology, where an extra step—to scan the slide once it is complete—is needed, FIBI produces an intrinsically digital image directly from the tissue with brief staining but without requiring additional processing. It may be possible, therefore, to develop a slide-free, non-destructive diagnostic work-flow for primary pathology diagnosis. Such a method promises improved speed, reduced cost, and better conservation of tissue for advanced ancillary studies.

## Data Availability

All data produced in the present study are available upon reasonable request to the authors.

## Acknowledgments

This work was supported in part by NIH grants 1R01EB028635, R33CA202881-01 and R21CA246359. SLT is supported in part by the National Center for Advancing Translational Sciences, National Institutes of Health, grant number UL1 TR001860.

## Conflict Disclosure

Histolix sponsored this study. RML and FF are co-founders and ADB is CMO of Histolix. TM has been an employee of Histolix. AMG is a consultant to Histolix. The other authors, with the exception of SLT, received payment from Histolix to perform the case review work but have no other relationship with the company.

## Supplemental Data

**Supplemental Table 1.**
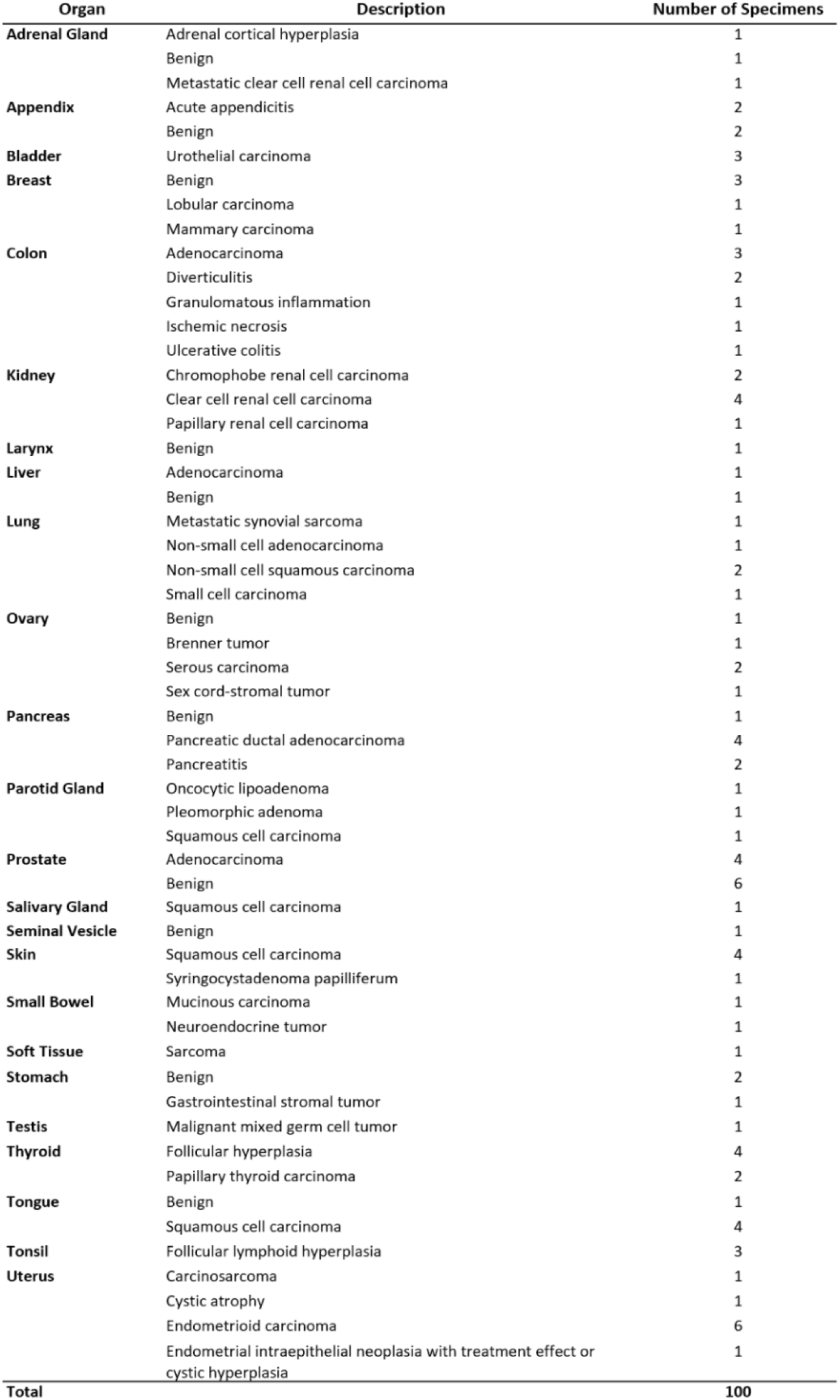
Distribution of tissues, reference diagnoses and number of cases in each category.

## Notes

### Funding Statement

Histolix, Inc. and NIH grants NIH 1R01EB028635, 
NIH R33CA20288, NIH R21 CA246359

### Author Declarations

The UC Davis IRB Administration reviewed the following protocol and determined that it was not deemed human subjects and IRB review was not required. FWA No: 00004557, Expiration Date: April 12, 2024, IORG: 0000251

## References

[1] R.D. Cardiff, J.P. Gregg, J.W. Miller, D.E. Axelrod, A.D. Borowsky, Histopathology as a predictive biomarker: strengths and limitations, The Journal of nutrition 136 (2006) 2673S–2675S.

[2] L.E. De Las Casas, D.G. Hicks, Pathologists at the Leading Edge of Optimizing the Tumor Tissue Journey for Diagnostic Accuracy and Molecular Testing, American journal of clinical pathology 155 (2021) 781–792.

[3] FDA allows marketing of first whole slide imaging system for digital pathology, Press Announcements, FDA, 2017.

[4] S. Mukhopadhyay, M.D. Feldman, E. Abels, R. Ashfaq, S. Beltaifa, N.G. Cacciabeve, H.P. Cathro, L. Cheng, K. Cooper, G.E. Dickey, Whole slide imaging versus microscopy for primary diagnosis in surgical pathology: a multicenter blinded randomized noninferiority study of 1992 cases (pivotal study), The American journal of surgical pathology 42 (2018) 39.

[5] A.D. Borowsky, E.F. Glassy, W.D. Wallace, N.S. Kallichanda, C.A. Behling, D.V. Miller, H.N. Oswal, R.M. Feddersen, O.R. Bakhtar, A.E. Mendoza, Digital Whole Slide Imaging Compared With Light Microscopy for Primary Diagnosis in Surgical PathologyA Multicenter, Double-Blinded, Randomized Study of 2045 Cases, Archives of pathology & laboratory medicine 144 (2020) 1245–1253.

[6] F. Fereidouni, Z.T. Harmany, M. Tian, A. Todd, J.A. Kintner, J.D. McPherson, A.D. Borowsky, M. Lechpammer, J. Bishop, S.G. Demos, R. Levenson, Microscopy with ultraviolet surface excitation for rapid slide-free histology, Nat Biomed Eng 1 (2017) 957–966.

[7] J.T. Liu, M.J. Mandella, J.M. Crawford, C.H. Contag, T.D. Wang, G.S. Kino, Efficient rejection of scattered light enables deep optical sectioning in turbid media with low-numerical-aperture optics in a dual-axis confocal architecture, Journal of biomedical optics 13 (2008) 034020.

[8] H. Nygren, B. Hagenhoff, P. Malmberg, M. Nilsson, K. Richter, Bioimaging TOF-SIMS: High resolution 3D imaging of single cells., Microscopy research and technique 70 (2007) 969–974.

[9] J.Q. Brown, A.B. Sholl, Applications of Structured Light Microscopy in Clinical Pathology, Microscopy Histopathology and Analytics, Optical Society of America, 2018, pp. MF1A. 1.

[10] M.L. Gabriele, G. Wollstein, H. Ishikawa, L. Kagemann, J. Xu, L.S. Folio, J.S. Schuman, Optical coherence tomography: history, current status, and laboratory work, Investigative ophthalmology & visual science 52 (2011) 2425–2436.

[11] M. Pekmezci, R.A. Morshed, P. Chunduru, B. Pandian, J. Young, J.E. Villanueva-Meyer, T. Tihan, E.A. Sloan, M.K. Aghi, A.M. Molinaro, Detection of glioma infiltration at the tumor margin using quantitative stimulated Raman scattering histology, Sci Rep 11 (2021) 1–11.

[12] J. Shi, T.T. Wong, Y. He, L. Li, R. Zhang, C.S. Yung, J. Hwang, K. Maslov, L.V. Wang, High-resolution, high-contrast mid-infrared imaging of fresh biological samples with ultraviolet-localized photoacoustic microscopy, Nat Photonics 13 (2019) 609–615.

[13] R. Lev, P.J. Stoward, On the use of eosin as a fluorescent dye to demonstrate mucous cells and other structures in tissue sections, Histochemie 20 (1969) 363–377.

[14] N.T. Goldsmith, Deep focus; a digital image processing technique to produce improved focal depth in light microscopy, Image Analysis & Stereology 19 (2000) 163–167.

[15] W. Jia, H. Zhang, X. He, Q. Wu, A comparison on histogram based image matching methods, Proceedings-IEEE International Conference on Video and Signal Based Surveillance 2006, AVSS 2006, 2006.

[16] T.W. Bauer, R.J. Slaw, Validating whole-slide imaging for consultation diagnoses in surgical pathology, Archives of Pathology and Laboratory Medicine 138 (2014) 1459–1465.

[17] A. Mukhopadhaya, J. Mendecki, X. Dong, L. Liu, S. Kalnicki, M. Garg, A. Alfieri, C. Guha, Localized hyperthermia combined with intratumoral dendritic cells induces systemic antitumor immunity, Cancer Res 67 (2007) 7798–7806.

[18] Y. Liu, R.M. Levenson, M.W. Jenkins, Slide over: advances in slide-free optical microscopy as drivers of diagnostic pathology, Am J Pathol (2021).

[19] M. Ragazzi, S. Piana, C. Longo, F. Castagnetti, M. Foroni, G. Ferrari, G. Gardini, G. Pellacani, Fluorescence confocal microscopy for pathologists, Mod Pathol 27 (2014) 460–471.

[20] Y.K. Tao, D. Shen, Y. Sheikine, O.O. Ahsen, H.H. Wang, D.B. Schmolze, N.B. Johnson, J.S. Brooker, A.E. Cable, J.L. Connolly, J.G. Fujimoto, Assessment of breast pathologies using nonlinear microscopy, Proc. Natl. Acad. Sci. 111 (2014) 15304--15309.

[21] M. Wang, H.Z. Kimbrell, A.B. Sholl, D.B. Tulman, K.N. Elfer, T.C. Schlichenmeyer, B.R. Lee, M. Lacey, J.Q. Brown, High-resolution rapid diagnostic imaging of whole prostate biopsies using video-rate fluorescence structured illumination microscopy, Cancer research 75 (2015) 4032–4041.

[22] C. Apelian, F. Harms, O. Thouvenin, A.C. Boccara, Dynamic full field optical coherence tomography: subcellular metabolic contrast revealed in tissues by interferometric signals temporal analysis, Biomedical optics express 7 (2016) 1511–1524.

[23] A.K. Glaser, N.P. Reder, Y. Chen, E.F. McCarty, C. Yin, L. Wei, Y. Wang, L.D. True, J.T. Liu, Light-sheet microscopy for slide-free non-destructive pathology of large clinical specimens, Nature biomedical engineering 1 (2017) 0084.

[24] Y. Liu, A.M. Rollins, M.W. Jenkins, CompassLSM: axially swept light-sheet microscopy made simple, Biomedical optics express 12 (2021) 6571–6589.

[25] S. You, H. Tu, E.J. Chaney, Y. Sun, Y. Zhao, A.J. Bower, Y.-Z. Liu, M. Marjanovic, S. Sinha, Y. Pu, Intravital imaging by simultaneous label-free autofluorescence-multiharmonic microscopy, Nature communications 9 (2018) 1–9.

